# LDCT-to-SDCT as a Bridge Problem: Single-Step Residual Endpoint Flow Matching for Real-Time Denoising

**DOI:** 10.64898/2026.08.27.26361520

**Authors:** K. Tomás de la Sotta, Jose M. Saavedra, Violeta Chang, Aline Xavier, Héctor Henríquez, Yessenia Orellana, Joaquín Curimil

## Abstract

Diffusion models achieve high reconstruction quality in lowdose computed tomography (LDCT), but their iterative sampling trajectories impose substantial computational costs. Unlike unconditional generation, paired LDCT reconstruction starts from an image that already contains the anatomy and spatial structure of the standard-dose CT (SDCT) target; reconstruction primarily requires correcting dose-related noise and artifacts. We therefore introduce Residual Endpoint Flow Matching (REFM), an LDCT reconstruction method that learns to transport an LDCT image directly toward its paired SDCT endpoint rather than defining a noise-to-image trajectory. REFM predicts the residual velocity along linear interpolations between both images and supports single-step and multi-step reconstruction using the same trained network. We evaluate five model capacities using 1 to 50 Euler steps against deterministic U-Net and diffusion-based baselines. Across all REFM variants, one-step inference consistently provides the highest reconstruction quality. On the TCIA validation set, REFM Base achieves 50.98 dB PSNR and 0.9865 SSIM at 94.54 fps, compared with 50.92 dB, 0.9847, and 9.26 fps for DDPM-10. REFM Small retains 50.71 dB while increasing throughput to 198.56 fps. Without fine-tuning, REFM Base also matches the 25-step DDPM baseline on the external Mayo Clinic dataset, although DDPM remains stronger on synthetically degraded CRLM images. Thus, our results show that exploiting paired anatomical correspondence enables diffusion-level LDCT reconstruction with a single step reconstruction.

## 1 Introduction

Computed tomography (CT) is central to diagnosis and clinical follow-up, but involves exposure to ionizing radiation. Low-dose CT (LDCT) protocols reduce this exposure at the cost of increased quantum noise and reconstruction artifacts, potentially obscuring low-contrast structures [1]. LDCT reconstruction therefore seeks to recover standard-dose CT (SDCT) image quality while preserving the anatomical information already present in the acquired image.

Many diffusion-based reconstruction methods recover the target through an iterative trajectory derived from a noise-based generative process. LDCT-specific adaptations can incorporate the observed image, but still rely on sequential sampling that limits reconstruction throughput. This formulation does not fully exploit paired LDCT–SDCT data: both images depict the same patient anatomy under aligned spatial geometry, so the task primarily requires correcting dose-related noise and artifacts rather than generating anatomical structure. We therefore introduce Residual Endpoint Flow Matching (REFM), an LDCT reconstruction method that learns the residual velocity along a direct linear path from each LDCT image to its paired SDCT target. This formulation replaces the noise-to-image trajectory with paired endpoint transport and supports both single-step and multi-step inference using the same network.

Our contributions are: (i) a paired conditional flow-matching formulation specifically designed for LDCT reconstruction; (ii) a systematic evaluation of one to 50 Euler steps across five model capacities; and (iii) diffusion-level reconstruction quality at substantially lower sampling cost. Experiments on TCIA show diffusion-level reconstruction quality with approximately 10× higher throughput than DDPM-10. We additionally evaluate performance across anatomical regions and generalization without fine-tuning on the independent Mayo Clinic and CRLM datasets.

The remainder of this paper is organized as follows. Section 2 reviews related methods, Section 3 presents REFM and the experimental protocol, Section 4 reports the results, and Section 5 presents final remarks.

## 2 Related Work

Deep learning methods initially addressed LDCT reconstruction through direct convolutional mappings [2], later incorporating adversarial, perceptual, and attention-based objectives [3, 4]. More recently, denoising diffusion probabilistic models have achieved strong performance in image restoration [5, 6] and LDCT reconstruction [7, 8]. Their iterative sampling process, however, requires repeated neural network evaluations. Accelerated samplers [9, 10] and trajectory-shortening strategies [11] reduce the number of network evaluations but remain sequential and multi-step, limiting their efficiency in volumetric and latency-sensitive applications.

Image-to-image bridge methods instead use degraded observations as informative source distributions. I^2^SB learns a stochastic bridge between degraded and clean image domains [12], while related work connects Schrödinger bridges with score and flow matching [13]. Flow Matching provides a deterministic alternative by learning the vector field of a prescribed probability path [14], and straight-path formulations such as Rectified Flow transport samples along interpolations between source and target endpoints [15]. Unlike stochastic image-to-image bridges, REFM defines a deterministic linear path between each aligned LDCT–SDCT pair and directly learns its endpoint residual. Our study further examines whether this paired transport can be reduced to a single network evaluation without sacrificing diffusion-level reconstruction quality.

## 3 Methodology

### 3.1 Residual Endpoint Flow Matching

Let *x*_*L*_ and *x*_*H*_ denote spatially aligned low-dose CT (LDCT) and standard-dose CT (SDCT) images, respectively. Residual Endpoint Flow Matching (REFM) directly transports *x*_*L*_ toward its paired target *x*_*H*_ through the linear conditional path

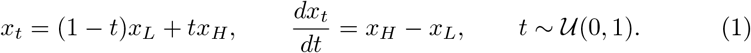

The conditional velocity is therefore the endpoint residual and remains constant along each paired interpolation. A time-conditioned network receives (*x*_*t*_, *t*) and learns this velocity by minimizing

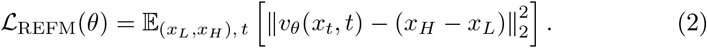

Unlike direct residual regression, which is supervised only at *x*_*L*_, REFM observes time-conditioned states throughout the paired path over successive training iterations.

Inference starts from *x*_0_ = *x*_*L*_ and uses *K* Euler steps:

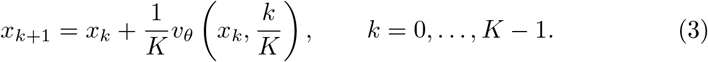

The reconstruction is 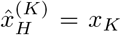. For *K* = 1, this reduces to 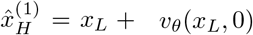. We evaluate the same trained network using *K* ∈ {1, 4, 8, 10, 25, 50} to determine whether numerical refinement improves reconstruction.

### 3.2 Model Family

All REFM variants use a time-conditioned U-Net operating on 256 × 256 single-channel images. Each network contains six encoder and decoder stages with symmetric skip connections and self-attention at one deep resolution and the bottleneck. Sinusoidal time embeddings, followed by a learned multilayer perceptron, condition every residual block. The five variants share this structure and differ in channel width and residual depth. Group normalization uses 32 groups except in Mini, which uses 16. Table 1 summarizes the architectures; each reported channel value represents two consecutive resolution stages.

**Table 1.** Architectural configurations of the REFM model family.

| Model | Params (M) | Channels | Layers/Block | GN Groups |
| --- | --- | --- | --- | --- |
| Base | 113.67 | 128–256–512 | 2 | 32 |
| Medium | 63.96 | 96–192–384 | 2 | 32 |
| Small | 43.53 | 96–192–384 | 1 | 32 |
| Mini | 30.24 | 80–160–320 | 1 | 16 |
| Tiny | 19.36 | 64–128–256 | 1 | 32 |

### 3.3 Experimental Setup

Models are trained using paired images from the TCIA Low-Dose CT and Projection dataset. The training partition contains 120 patients: 45 chest, 36 brain, and 39 liver. The independent validation partition contains 30 patients: 5 chest, 14 brain, and 11 liver.

External generalization is evaluated without fine-tuning on ten paired quarter-dose and standard-dose cases from the Mayo Clinic Low-Dose CT Grand Challenge and on 96 patients from the Colorectal Liver Metastases (CRLM) dataset. For CRLM, synthetic LDCT images are generated at dose settings 1*/*2, 1*/*4, and 1*/*8 using the Poisson–Gaussian noise model and preprocessing protocol from our previous benchmark. All methods operate on single-channel 256 × 256 images and share the same evaluation pipeline within each dataset.

Baselines comprise a conventional U-Net trained to reconstruct SDCT directly from LDCT and a conditional DDPM trained on the same TCIA partition. DDPM is evaluated using *K* ∈ {10, 25, 150, 500, 1000}reverse-sampling steps. Reconstruction quality is reported as image-level mean ± standard deviation using PSNR and SSIM. Sampler throughput is measured in images per second, excluding data loading, disk access, and metric computation, using the same hardware and timing procedure for all methods.

## 4 Results

### 4.1 One-step inference and model scaling

Across all evaluated capacities, REFM achieved its highest reconstruction quality using one Euler step. REFM Base reached 50.98 dB and 0.9865 SSIM at 94.54 fps, compared with 50.92 dB, 0.9847, and 9.26 fps for DDPM-10, providing comparable quality at approximately 10.2× higher throughput. Increasing REFM Base from one to 50 steps reduced PSNR to 50.05 dB and throughput to 1.86 fps, with the same overall trend observed across all variants.

DDPM also saturated early: increasing its trajectory from 10 to 1000 steps changed PSNR by only 0.20 dB while reducing throughput from 9.26 to 0.09 fps. Thus, DDPM-10 provides its strongest quality–efficiency trade-off, whereas numerical refinement provides no benefit for the evaluated REFM models.

**Fig 1.**
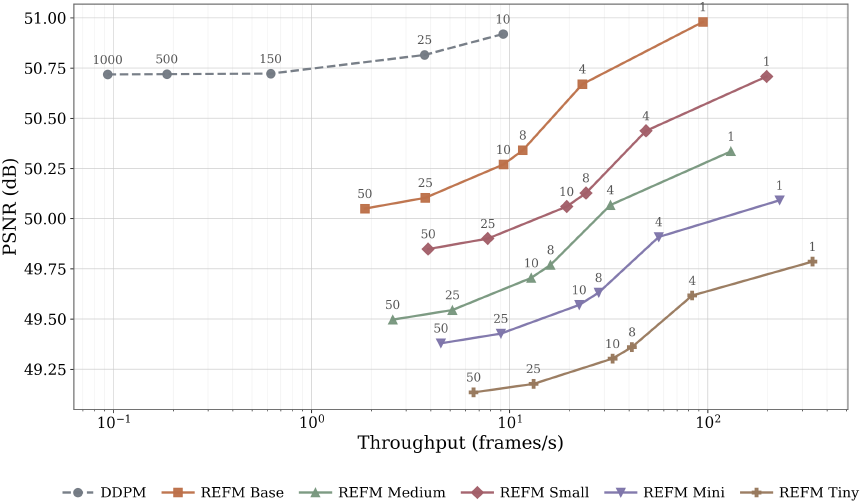
PSNR–throughput trade-off on TCIA. Labels indicate the number of integration or sampling steps. REFM achieves its highest reconstruction quality using one Euler step.

Table 2 summarizes the one-step results. Base achieved the highest quality, whereas Tiny reached 338.02 fps with a 1.19 dB reduction. Small provided the strongest intermediate trade-off, remaining 0.27 dB below Base while operating approximately 2.1× faster. Performance was not strictly monotonic with capacity, as Small outperformed the larger Medium model.

**Table 2.**
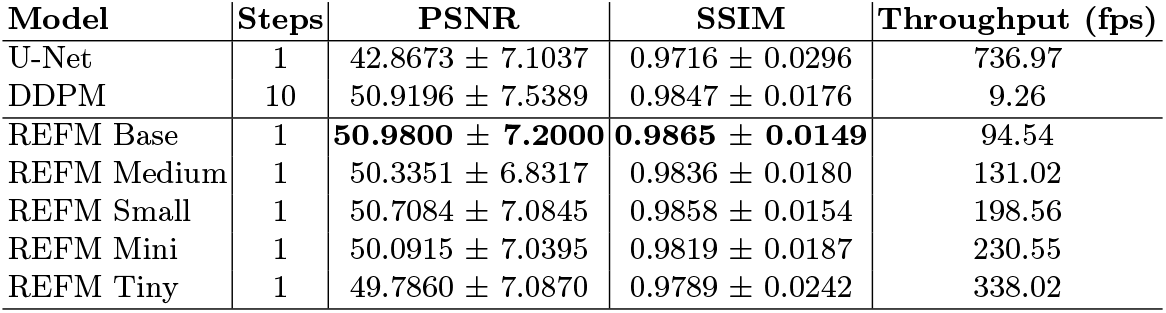
Performance comparison on the TCIA validation dataset. All REFM variants use one-step inference.

### 4.2 Anatomical performance

Table 3 reports reconstruction performance separately for liver, chest, and brain images. Substantial differences were observed across anatomical subsets. Using one-step REFM Base, PSNR reached 54.74 dB for liver, 43.68 dB for chest, and 63.53 dB for brain. The corresponding SSIM values were 0.9975, 0.9715, and 0.9996, respectively. Chest therefore represented the most challenging subset for all evaluated methods.

**Table 3.** TCIA performance for liver (*N* = 11), chest (*N* = 5), and brain (*N* = 14). REFM uses one step.

| Model | Steps | Liver |  | Chest |  | Brain |  |
| --- | --- | --- | --- | --- | --- | --- | --- |
|  |  | PSNR | SSIM | PSNR | SSIM | PSNR | SSIM |
| U-Net | 1 | 47.3014 ± 2.2580 | 0.9937 ± 0.0020 | 35.3807 ± 1.7669 | 0.9414 ± 0.0200 | 53.5157 ± 2.1026 | 0.9989 ± 0.0004 |
| DDPM | 10 | **54.9467 ± 1.6594** | **0.9976 ± 0.0009** | 43.2048 ± 1.6076 | 0.9673 ± 0.0135 | **63.9944 ± 3.6188** | **0.9996 ± 0.0003** |
| REFM Base | 1 | 54.7394 ± 1.8218 | 0.9975 ± 0.0009 | **43.6774 ± 1.4895** | **0.9715 ± 0.0107** | 63.5297 ± 3.6519 | **0.9996 ± 0.0003** |
| REFM Medium | 1 | 54.0159 ± 2.3901 | 0.9940 ± 0.0126 | 43.4080 ± 1.5342 | 0.9686 ± 0.0133 | 61.8234 ± 2.7072 | 0.9992 ± 0.0003 |
| REFM Small | 1 | 54.5010 ± 1.9252 | 0.9971 ± 0.0025 | 43.4719 ± 1.4877 | 0.9704 ± 0.0109 | 62.9144 ± 3.0542 | 0.9995 ± 0.0003 |
| REFM Mini | 1 | 53.7182 ± 2.4787 | 0.9930 ± 0.0130 | 43.0516 ± 1.4769 | 0.9657 ± 0.0126 | 62.2094 ± 3.4500 | 0.9990 ± 0.0005 |
| REFM Tiny | 1 | 53.8162 ± 1.9143 | 0.9944 ± 0.0056 | 42.9583 ± 1.5205 | 0.9605 ± 0.0174 | 61.8738 ± 3.4321 | 0.9993 ± 0.0006 |

The comparison with DDPM-10 varied by anatomical region. REFM Base produced higher performance on chest images, improving PSNR from 43.20 to 43.68 dB and SSIM from 0.9673 to 0.9715. DDPM-10 achieved slightly higher PSNR on liver and brain, although SSIM remained nearly identical between both approaches.

Additional REFM integration steps did not provide a consistent improvement in any anatomical subset. The largest degradation was observed for chest images, whereas liver and brain results remained comparatively stable. These findings show that the advantage of one-step inference is preserved across the evaluated anatomical regions. Figure 2 shows consistent noise suppression across the REFM family, with reconstructions visually comparable to DDPM despite using a single network evaluation.

**Fig 2.**
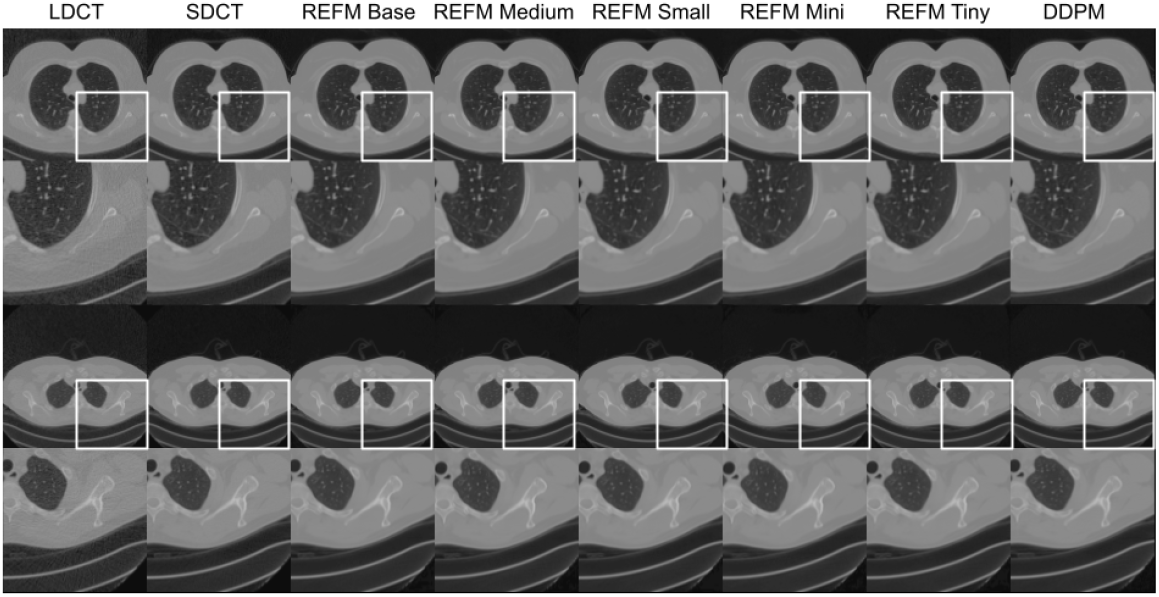
Two representative TCIA cases with enlarged regions (white boxes). REFM uses one Euler step and DDPM 10 reverse steps; all images share the same intensity window.

### 4.3 Generalization to external datasets

To evaluate generalization beyond the TCIA validation distribution, all models were evaluated without retraining or fine-tuning on the Mayo Clinic and CRLM datasets. Mayo provides paired quarter-dose and standard-dose CT images, whereas the CRLM experiments use synthetically generated LDCT inputs at three dose settings.

**Table 4.** Comparison between the best DDPM baseline and the proposed one-step REFM variants on the CRLM dataset (*N* = 96) under three simulated dose-reduction settings: 1/2, 1/4, and 1/8. DDPM requires 25 sampling steps, whereas all REFM variants perform reconstruction in a single step.

| Model | Steps | 1/2 |  | 1/4 |  | 1/8 |  |
| --- | --- | --- | --- | --- | --- | --- | --- |
|  |  | PSNR | SSIM | PSNR | SSIM | PSNR | SSIM |
| UNet | | 28.5707 ± 4.4288 | 0.8893 ± 0.0664 | 29.0268 ± 4.4727 | 0.8938 ± 0.0669 | 29.5138 ± 4.4448 | 0.8940 ± 0.0698 |
| DDPM | 25 | 35.3986 ± 3.7807 | 0.9061 ± 0.0505 | 36.1279 ± 3.8794 | 0.9191 ± 0.0496 | 36.9123 ± 3.7570 | 0.9303 ± 0.0507 |
| REFM Base | 1 | 33.8684 ± 4.7908 | 0.8778 ± 0.0736 | 34.3378 ± 4.9529 | 0.8838 ± 0.0753 | 34.9610 ± 5.0536 | 0.8921 ± 0.0780 |
| REFM Medium | 1 | 34.0936 ± 4.7867 | 0.8789 ± 0.0774 | 34.3527 ± 4.9153 | 0.8776 ± 0.0802 | 34.6107 ± 5.0358 | 0.8741 ± 0.0848 |
| REFM Small | 1 | 34.6668 ± 4.8055 | 0.9048 ± 0.0727 | 34.9976 ± 4.9192 | 0.9045 ± 0.0753 | 35.3421 ± 5.0141 | 0.9015 ± 0.0799 |
| REFM Mini | 1 | 34.2351 ± 4.8520 | 0.8813 ± 0.0794 | 34.5154 ± 4.9754 | 0.8803 ± 0.0820 | 34.8028 ± 5.0986 | 0.8773 ± 0.0865 |
| REFM Tiny | 1 | 34.3386 ± 4.6956 | 0.8938 ± 0.0760 | 34.6209 ± 4.8296 | 0.8924 ± 0.0791 | 34.9066 ± 4.9544 | 0.8885 ± 0.0837 |

**Table 5.** Comparison between the best DDPM baseline and the proposed one-step REFM variants on the MAYO Grand Challenge 2016 validation dataset (*N* = 10).

| Model | Steps | PSNR | SSIM |
| --- | --- | --- | --- |
| UNet | | 42.2123 ± 3.1468 | 0.9814 ± 0.0107 |
| DDPM | 25 | 48.8271 ± 2.8373 | 0.9861 ± 0.0082 |
| REFM Base | 1 | 48.8278 ± 2.8393 | 0.9866 ± 0.0073 |
| REFM Medium | 1 | 47.3676 ± 3.2844 | 0.9728 ± 0.0197 |
| REFM Small | 1 | 48.4131 ± 2.9489 | 0.9834 ± 0.0102 |
| REFM Mini | 1 | 47.3585 ± 3.3762 | 0.9721 ± 0.0219 |
| REFM Tiny | 1 | 47.4512 ± 3.3437 | 0.9729 ± 0.0211 |

On the Mayo Clinic dataset, one-step REFM Base matched the reconstruction quality of DDPM-25. REFM Base reached 48.8278 dB and an SSIM of 0.9866, compared with 48.8271 dB and 0.9861 for DDPM. The observed differences are negligible in magnitude, indicating effectively equivalent reconstruction performance. REFM nevertheless requires only one network evaluation, compared with 25 reverse-sampling steps for DDPM.

The smaller REFM variants exhibited a larger reduction on Mayo. REFM Small achieved 48.41 dB, remaining 0.41 dB below Base, whereas Medium, Mini, and Tiny reached approximately 47.4 dB. This indicates that the external distribution is more sensitive to model configuration than the TCIA validation set.

On CRLM, DDPM-25 achieved the highest reconstruction quality across the three simulated dose settings. Among the proposed models, REFM Small provided the most consistent external performance, reaching 34.67, 35.00, and 35.34 dB at the 1*/*2, 1*/*4, and 1*/*8 settings, respectively. Its corresponding SSIM values were also higher than those of the remaining REFM variants. REFM therefore did not fully preserve the in-distribution performance of DDPM on CRLM, although it reduced the reconstruction trajectory from 25 steps to one.

Taken together, the external evaluations reveal two distinct behaviors. REFM Base matched DDPM on paired Mayo data, whereas a moderate performance gap remained on synthetically degraded CRLM images. These results support the ability of one-step REFM to generalize beyond TCIA, while also showing that its relative performance depends on the external data distribution and degradation process.

### 4.4 An Overall Discussion

The results show that paired LDCT–SDCT reconstruction can achieve diffusion-level quality without a long noise-to-image trajectory. REFM does not substantially improve peak reconstruction quality; rather, it preserves it at approximately one tenth of the DDPM-10 sampling cost. This behavior was consistent across all capacities, with one Euler step providing the highest quality and throughput.

The degradation with additional REFM steps does not imply that the learned field is exactly constant. Repeated evaluations may instead accumulate errors or produce states that differ from the paired interpolations observed during training. One-step inference avoids this effect by estimating the complete endpoint displacement directly from LDCT. Although this update resembles residual prediction, REFM is trained on time-conditioned intermediate states. Since the U-Net baseline is not capacity-matched, the results establish REFM as an effective one-step formulation without isolating the contribution of intermediate-state supervision.

Model scaling produced a non-monotonic quality–speed trade-off. Base achieved the highest quality, while Tiny reached 338.02 fps with a 1.19 dB reduction. Small remained only 0.27 dB below Base while operating approximately 2.1× faster, but outperformed the larger Medium model. Anatomical differences may reflect acquisition and intensity characteristics in addition to anatomical complexity.

External evaluation revealed distinct generalization behaviors. One-step REFM Base matched DDPM-25 on paired Mayo data without fine-tuning, whereas DDPM retained an advantage on synthetically degraded CRLM images and Small generalized better than Base. Direct paired transport may therefore be sensitive to changes in the degradation process. Moreover, the reported throughput measures sampler efficiency rather than end-to-end clinical latency, since preprocessing, data transfer, and volume assembly are excluded.

Several limitations remain. PSNR and SSIM do not measure diagnostic fidelity, and image-level statistics ignore within-patient correlation. Patient-level, task-based, and volumetric evaluation remains necessary. Within these limitations, REFM preserves diffusion-level reconstruction quality using a single network evaluation.

## 5 Conclusions

Reducing CT radiation without compromising image quality remains clinically important. REFM addresses this challenge through direct residual transport, matching DDPM-level quality on TCIA and Mayo with a single network evaluation and substantially higher throughput. These results represent a promising step toward efficient, high-fidelity LDCT reconstruction in latency-sensitive settings.

## Data Availability

All data produced in the present study are available upon reasonable request to the authors

